# Association Between Nursing Home Crowding and COVID-19 Infection and Mortality in Ontario, Canada

**DOI:** 10.1101/2020.06.23.20137729

**Authors:** Kevin A. Brown, Aaron Jones, Nick Daneman, Adrienne K. Chan, Kevin L. Schwartz, Gary E. Garber, Andrew P. Costa, Nathan M. Stall

## Abstract

**Importance:** Nursing home residents have been disproportionately impacted by the COVID-19 epidemic. Prevention recommendations have emphasized frequent testing of healthcare personnel and residents, but additional strategies are needed to protect nursing home residents.

**Objective:** We developed a reproducible index of nursing home crowding and determined whether crowding was associated with incidence of COVID-19 in the first months of the COVID-19 epidemic.

**Design, Setting, and Participants:** Population-based retrospective cohort study of over 78,000 residents of 618 distinct nursing homes in Ontario, Canada from March 29 to May 20, 2020.

**Exposure:** The nursing home crowding index equalled the average number of residents per bedroom and bathroom.

**Outcomes:** Primary outcomes included the cumulative incidence of COVID-19 infection and mortality, per 100 residents; introduction of COVID-19 into a home (≥1 resident case) was a negative tracer.

**Results:** Of 623 homes in Ontario, we obtained complete information on 618 homes (99%) housing 78,607 residents. A total of 5,218 residents (6.6%) developed COVID-19 infection, and 1,452 (1.8%) died with COVID-19 infection as of May 20, 2020. COVID-19 infection was distributed unevenly across nursing homes: 4,496 (86%) of infections occurred in just 63 (10%) of homes. The crowding index ranged across homes from 1.3 (mainly single-occupancy rooms) to 4.0 (exclusively quadruple occupancy rooms); 308 (50%) homes had high crowding index (≥2). Incidence in high crowding index homes was 9.7%, versus 4.5% in low crowding index homes (p<0.001), while COVID-19 mortality was 2.7%, versus 1.3%. The likelihood of COVID-19 introduction did not differ (31.3% vs 30.2%, p=0.79). After adjustment for regional, nursing home, and resident covariates, the crowding index remained associated with increased risk of infection (RR=1.72, 95% Confidence Interval [CI]: 1.11-2.65) and mortality (RR=1.72, 95%CI: 1.03-2.86). Propensity score analysis yielded similar conclusions for infection (RR=2.06, 95%CI: 1.34-3.17) and mortality (RR=2.09, 95%CI: 1.30-3.38). Simulations suggested that converting all 4-bed rooms to 2-bed rooms would have averted 988 (18.9%) infections of COVID-19 and 271 (18.7%) deaths.

**Conclusions and Relevance:** Crowding was associated with higher incidence of COVID-19 infection and mortality. Reducing crowding in nursing homes could prevent future COVID-19 mortality.

## Background

Nursing home residents have suffered the brunt of the Coronavirus Disease 2019 (COVID-19) epidemic. Recent estimates suggest that nursing home residents account for approximately 35% of COVID-19 deaths in the US, and between 66% and 81% in Canada.^1,2^ Compared to community dwelling older adults, nursing home residents are 5 times more likely to die with COVID-19.^3^ The concentration of COVID-19 mortality among nursing home residents is due to a combination of: 1) high risk of contracting infection due to congregant living, exposure to staff, and inability to physically distance and practice hand hygiene as a result of cognitive and functional impairment, and 2) high mortality due to advanced age (immunosenescence) and multimorbidity.^4–6^

While factors associated with COVID-19 complications are well documented, it is more difficult to understand why COVID-19 may impact some nursing homes but spare others, and what policy makers can do to prevent further mortality among highly vulnerable nursing home residents. Incidence in the region surrounding a nursing home and nursing home size drive the probability of introduction of COVID-19, by visitors, or staff.^3^ Once introduced, the high prevalence of asymptomatic infection and atypical presentation among nursing home residents,^7^ along with an important pre-symptomatic period, enable the virus to spread undetected, and may have thwarted efforts at infection control focused on symptomatic individuals.^8^ Past studies suggest that crowded nursing homes may be more susceptible to influenza outbreaks, and associations observed with domestic crowding would suggest plausibility for COVID-19 as well.^9–11^ A recent study demonstrated that for-profit nursing homes in Ontario had larger COVID-19 outbreaks compared to municipally run homes, and were less likely to meet current design standards.^12^

We examined the association between nursing home crowding and COVID-19 incidence across the entire nursing home system of Ontario, Canada (>78,000 nursing home residents, >600 nursing homes), during the first months of the COVID-19 pandemic in Ontario. Specifically, we sought to: 1) develop a replicable nursing home crowding index; 2) test the association of the crowding index with the incidence of COVID-19 and the number of resident deaths; and 3) estimate the probable number of preventable infections and deaths by interventions focused on reducing crowding in nursing homes.

## Methods

### Ethics Statement

The study was approved by the Research Ethics Board of the University of Toronto. The board waived the need for consent because there was no contact with nursing home residents and anonymity was assured.

### Study Design and Data Sources

We conducted a retrospective cohort study across all nursing homes in Canada’s most populous province of Ontario from March 29, 2020, the date of the first reported Ontario nursing home outbreak until May 20, 2020 (52 days). Data used for this study were obtained from the Ontario Ministries of Health and Long-Term Care as part of the province’s Emergency Modeling Table and were derived from 4 sources: 1) information on characteristics of nursing homes from the Ontario Ministry of Long-Term Care’s Inspections Branch, as well as 2) their COVID Tracking Tool, which includes data that has been specifically collected to measure home-level COVID-19 infections and deaths among nursing home residents, 3) the integrated Public Health Information System, which was used to identify incidence of COVID-19 in the 35 public health regions surrounding nursing homes, and 4) the Canadian Census population estimate (2020), for information on community size and health region population.^13^

### Exposure – Nursing Home Crowding Index

The nursing home crowding index was defined as the average number of occupants per room and bathroom across an entire home, following the equation: N_residents_ ÷ (½N_bedrooms_ + ½N_bathrooms_). This translated to weights per resident according to the room they occupied: single occupancy room with private bathroom – 1; single occupancy room with a shared bathroom – 1.5; double occupancy room (with shared bathroom) – 2; and quadruple occupancy room – 4. In Ontario nursing homes, there are no rooms with a maximum occupancy of 3 or greater than 4. We defined homes with crowding index ≥2 as high crowding index homes, and homes <2 as low. When reporting risk ratios for models with the linear crowding index, we compared estimates for homes with an index of 1.5 (an equal mix of single- and double occupancy rooms), to homes with an index of 3 (an equal mix of double- and quadruple occupancy rooms).

### Outcomes

The primary outcomes were the cumulative incidence of COVID-19 per 100 nursing home residents and the COVID-19 associated deaths per 100 nursing home residents. To identify potential confounding, we also examined COVID-19 introduction (defined as ≥1 confirmed COVID-19 resident case in a nursing home) as a negative tracer outcome, since crowding should only impact COVID-19 transmission within nursing homes, rather than risk of importation.^14^

### Nursing Home Characteristics

In addition to the crowding index, we examined 4 home design features, including: (1) the size of the facility (<100, ≥100 beds), (2) the type of ownership of the facility (private for-profit, private non-profit, or municipal), (3) the proportion of 1-bed, 2-bed, and 4-bed rooms, (4) the design standard (pre-1999 versus 1999 or later) that each home met.^15^ Prior to 1999, there was no universal design standard in the province, and as such the design of homes was heterogeneous. Changes to the standard since 1999 have been incremental.^16^ In terms of design elements of particular relevance for infection control, the 1999 design standard specifies: (1) that all homes have separate and independent sub-units with a maximum size of 40 occupants, known as home areas, having separate entrances, and independent common and dining areas, and (2) that resident rooms have no more than 2 beds.

### Regional Characteristics

We examined the incidence of COVID-19 in the surrounding public health region (N=35), and excluding nursing home infections, per 10,000 population, and community size (<10,000, 10,000 to 499,999, ≥500,000 individuals). Health region population and community population size were based on Statistics Canada projections for 2020.^13^

### Resident Characteristics

Four different resident characteristics, measured at the home level, were obtained from the most recent Resident Assessment Instrument Minimum Dataset (RAI-MDS),^17^ from August 2019: female sex (%), age ≥85 years (%), dementia (%), and the mean of the activities of daily living (ADL) impairment scale (0: no impairment, to 28: complete impairment).

### Statistical Analysis

All reported p-values were based on 2-sided testing. Model-based estimates reported 95% confidence intervals (CIs). All analyses were conducted using SAS software (version 9.4).

We developed unadjusted and adjusted regression models for each nursing home outcome. For all analyses, we included region-level random effects to account for clustering of incidence in regions.^18^ Quasi-Poisson regression was used to model count outcomes, using the logarithm of the number of beds in the home as an offset, while logistic regression was used to model COVID-19 introduction. Unadjusted models included just the region-level random effect. Adjusted models included the crowding index, nursing home size, and ownership, along with the 4 resident and 2 region covariates.

We used propensity score matching to adjust for potential confounding between the crowding index and resident or regional factors.^19^ The propensity score was the predicted probability for a home to have a high crowding index based on a logistic regression model that included nursing home size, and ownership, along with the 4 resident and 2 region covariates and random-intercepts for region. Homes with a high crowding index were matched 1:1 to homes with a low crowding index within a caliper of 0.2 of the logit of the propensity score, using a greedy matching algorithm.^20^ We then assessed the balance using standardized differences and considered >10% a meaningful difference.^20^ Quasi-Poisson regression, with random effects corresponding to matched pairs, was then used to estimate the effects of the crowding index.

We used a simulation to estimate the proportion of COVID-19 infections and deaths that could have been averted if nursing homes were less crowded.^21^ Specifically, we simulated scenarios where: 1) all 4-bed rooms were replaced with 2-bed rooms, and 2) all 2- and 4-bed rooms were replaced with 1-bed rooms. These scenarios were estimated by modifying the underlying crowding indexes of homes; for instance, if a home had 40% 1-bed, 30% 2-bed and 30% 4-bed rooms (crowding index=2.2), we estimated incidence as if the home had 40% 1-bed and 60% 2-bed rooms (crowding index=1.6), then as if the home was fully 1-bed rooms (crowding index=1.0). We then used regression coefficients from the adjusted regression model, to estimate incidence for all homes under these 2 simulated interventions. The preventable fraction was the simulated incidence divided by the incidence observed.

## Results

### Cohort

Of Ontario’s 623 homes, we were able to obtain complete information on 618 homes (99%); information on room types was missing for 5 homes. There were 78,607 residents in these homes at the time of the first case in March 29, 2020. Across the homes, 55% of residents were 85 or older, 69% were female, 70% had dementia and the average ADL score was 17.5 (Table 1).

**Table 1.** Characteristics of low, and high crowding index nursing homes. Presented as N (% of homes) or median (IQR).

|  | All Homes | Crowding Index |  | p |
| --- | --- | --- | --- | --- |
|  |  | Low<br>1.3 to 1.9 | High<br>2.0 to 4.0 |  |
| Homes | 618 (100) | 310 (50.1) | 308 (49.9) |  |
| Resident characteristics |  |  |  |  |
| Female (%) | 69.2 (64.6, 73.7) | 70.5 (67.3, 73.9) | 67.3 (61.9, 73.4) | <0.001 |
| Age ≥85 (%) | 55.3 (48.7, 61.0) | 58.3 (54.3, 63.1) | 50.4 (43.9, 58.1) | <0.001 |
| Dementia (%) | 70.0 (63.0, 76.7) | 72.0 (66.3, 78.0) | 67.1 (58.8, 75.2) | <0.001 |
| ADL Impairment Scale (mean) | 17.7 (16.0, 19.0) | 18.0 (16.4, 19.1) | 17.6 (15.8, 18.8) | 0.028 |
| Region characteristics |  |  |  |  |
| Regional COVID-19 incidence<br>(per 10,000 residents) |  |  |  |  |
| 1<sup>st</sup> Quartile (0.11, 0.46) | 148 (24.0) | 58 (18.7) | 90 (29.2) | 0.022 |
| 2<sup>nd</sup> Quartile (0.51, 0.93) | 141 (22.8) | 74 (23.4) | 67 (21.8) |  |
| 3<sup>rd</sup> Quartile (0.94, 1.29) | 159 (25.7) | 88 (28.4) | 71 (23.1) |  |
| 4<sup>th</sup> Quartile (1.30, 1.88) | 170 (27.5) | 90 (29.0) | 80 (26.0) |  |
| Community population size |  |  |  |  |
| ≥500,000+ | 253 (40.9) | 146 (47.1) | 107 (34.7) | <0.001 |
| 10,000-499,999 | 224 (36.3) | 116 (37.4) | 108 (35.1) |  |
| <10,000 | 141 (22.8) | 48 (15.5) | 93 (30.2) |  |
| Nursing home characteristics |  |  |  |  |
| Ownership (N, %) |  |  |  |  |
| Private for-profit | 358 (57.9) | 127 (41.0) | 231 (75.0) | <0.001 |
| Private non-profit | 159 (25.7) | 101 (32.6) | 58 (18.8) |  |
| Municipal | 101 (16.3) | 82 (26.5) | 19 (6.2) |  |
| Design standard (N, %) |  |  |  |  |
| 1999 | 310 (50.2) | 310 (100) | 0 (0) | <0.001 |
| Pre-1999 | 308 (49.8) | 0 (0) | 308 (100) |  |
| Size of home (N, %) |  |  |  |  |
| <100 | 244 (39.5) | 69 (22.3) | 175 (56.8) | <0.001 |
| ≥100 | 374 (60.5) | 241 (77.7) | 133 (43.2) |  |
| Room occupancy |  |  |  |  |
| Single (%) | 45.0 (13.0, 60.0) | 59.9 (59.0, 60.0) | 13.1 (6.5, 22.2) | <0.001 |
| Double (%) | 40.0 (33.1, 41.2) | 40.1 (40.0, 41.0) | 33.3 (21.7, 41.8) | <0.001 |
| Quadruple (%) | 0 (0, 50.0) | 0 (0, 0) | 50.2 (40.3, 57.1) | <0.001 |
| Crowding index | 2.0 (1.4, 2.9) | 1.4 (1.4-1.5) | 2.9 (2.7, 3.1) | <0.001 |
| Outcomes |  |  |  |  |
| Residents | 78,607 | 46,028 | 32,579 |  |
| Infections (N, % residents) | 5,218 (6.64) | 2,071 (4.50) | 3,147 (9.66) | <0.001 |
| Mortality (N, % residents) | 1,452 (1.85) | 578 (1.26) | 874 (2.68) | <0.001 |
| ≥1 Infection (N, %) | 190 (30.7) | 97 (31.3) | 83 (30.2) | 0.79 |

Abbreviations: ADL, activities of daily living; IQR, interquartile range; Q, quartile.

### Nursing Homes Characteristics

Across the province 36.9%, 37.3%, and 25.8% of residents were located in single, double and quadruple-bedded rooms, respectively. Half (49.8%) of nursing homes were built according to a pre-1999 design standard that allowed quadruple-bedded rooms. The nursing home crowding index varied from 1.3 to 4.0 (Figure 1). Homes meeting the 1999 design standard had a mix of single and double occupancy rooms and a crowding index between 1.3 and 1.9, while homes with older design standards had a mix of single, double and quadruple occupancy rooms, and crowding indexes that ranged between 2.0 and 4.0.

**Figure 1.**
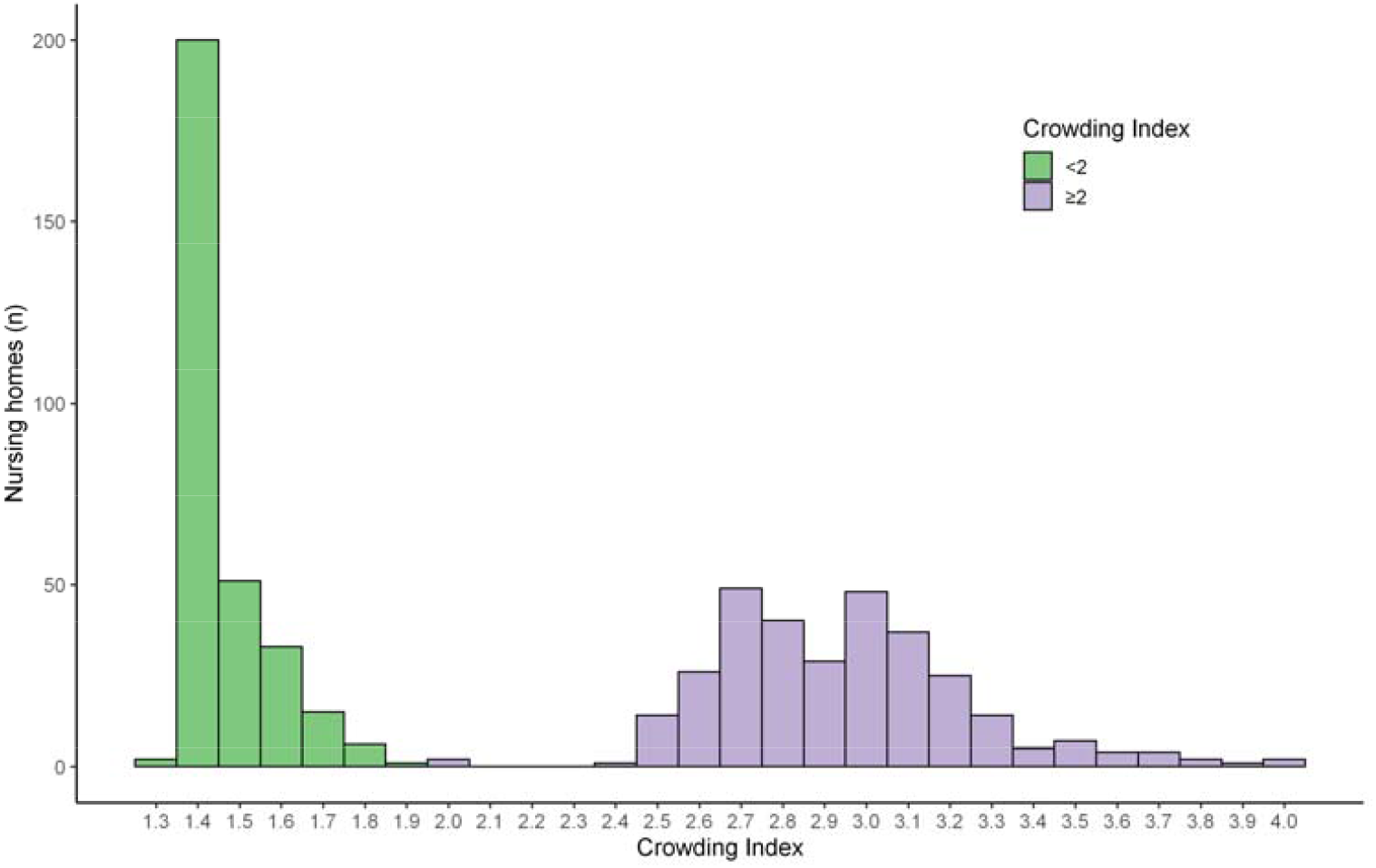
Nursing home crowding index, across 618 homes in Ontario, Canada

### Outcomes

Of 78,607 residents, 5,218 (6.6%) developed COVID-19 infection, and 1,452 (1.8%) died with COVID-19 infection as of May 20, 2020. The case fatality was 27.8%. COVID-19 infection was distributed unequally across nursing homes in the province, with 4,496 (86%) of infections occurring in just 63 (10%) of homes.

### Association Between Crowding Index and COVID-19 Incidence and Mortality

COVID-19 incidence in high crowding index homes was 9.7%, compared to 4.5% in low crowding index homes (p<0.001). Similarly, mortality was 2.7% in high crowding index homes, versus 1.3% in low crowding index homes (p<0.001). The likelihood of an outbreak occurring did not differ according to the crowding index (31.3% versus 30.2%, p=0.79). Outbreaks in crowded nursing homes tended to be larger: there were 9 outbreaks involving more than 100 residents in high crowding homes, compared to just 1 in low crowding homes (Figure 2).

**Figure 2.**
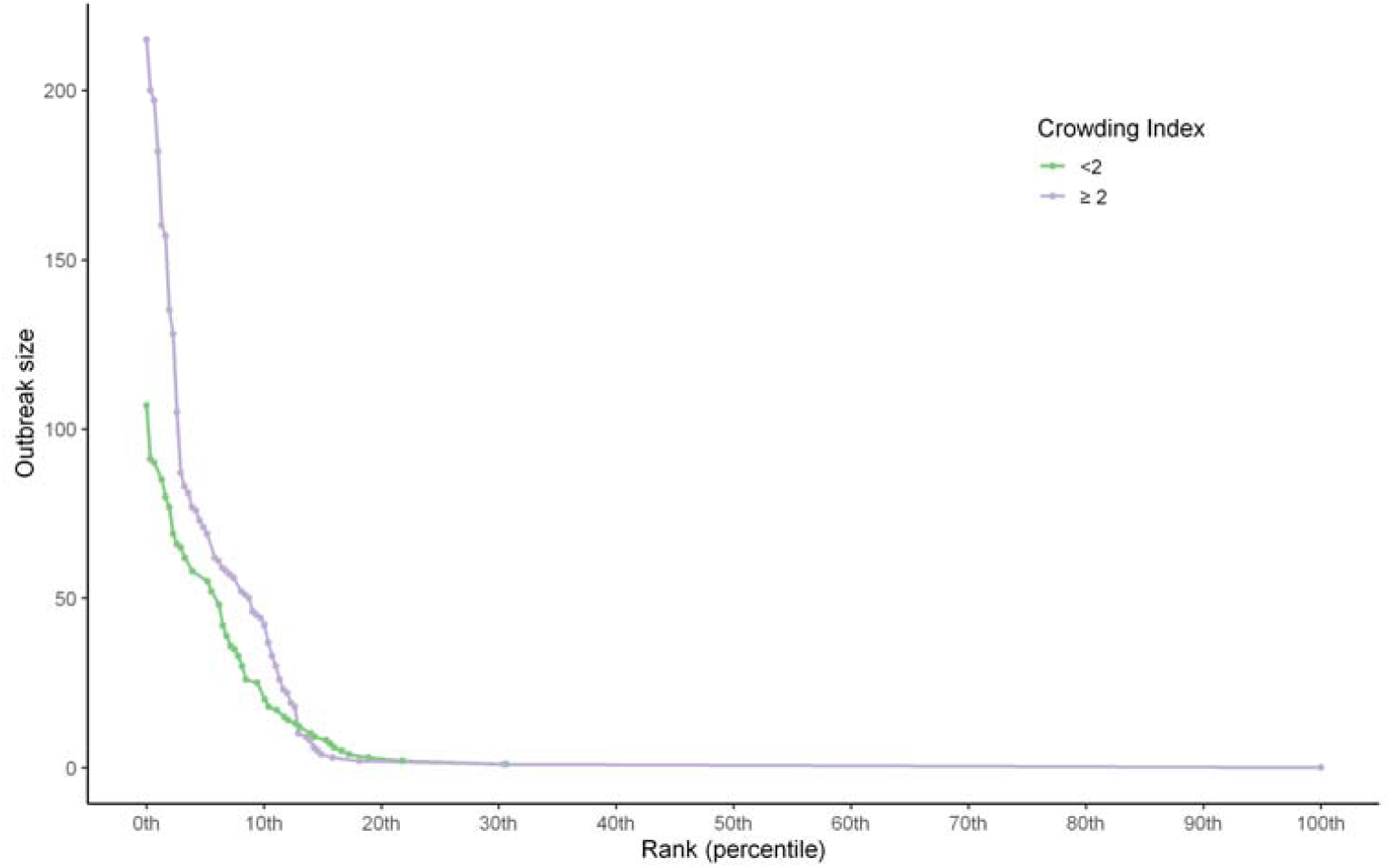
Ranked COVID-19 outbreak size across 618 nursing homes in Ontario, Canada.

Regression models based on the linear crowding index (Figure 3, Table 2), suggested that, compared to a home with an index of 1.5, homes with an index of 3 had double the COVID-19 incidence (relative risk [RR]=2.05, 95%CI: 1.49-2.70). After adjustment, the crowding index remained associated with increased risk (RR=1.83, 95%CI: 1.25-2.70). The crowding index was similarly associated with COVID-19 mortality (unadjusted RR=1.97, 95%CI: 1.36-2.84; adjusted RR=1.76, 95%CI: 1.13-2.76). The crowding index was not associated with the probability of COVID-19 being introduced (unadjusted OR=1.13, 95%CI: 0.70-1.84; adjusted OR=1.45 95%CI: 0.95-2.19), but other factors, including regional incidence, and nursing home size, were.

**Table 2.** Nursing home characteristics and incidence of COVID-19 infections (per 100 residents) and mortality (per 100 residents)

|  | COVID-19 Incidence<br>RR (95% CI) |  | COVID-19 Mortality<br>RR (95% CI) |  | COVID-19 Introduction (≥1 Infection)<br>OR (95% CI) |  |
| --- | --- | --- | --- | --- | --- | --- |
|  | Unadjusted | Adjusted | Unadjusted | Adjusted | Unadjusted | Adjusted |
| **Regional factors** |  |  |  |  |  |  |
| Regional COVID-19 incidence<br>(per 10,000 residents) |  |  |  |  |  |  |
| 1<sup>st</sup> Quartile (0.11, 0.46) | Reference | Reference | Reference | Reference | Reference | Reference |
| 2<sup>nd</sup> Quartile (0.51, 0.93) | 1.79 (0.37-8.77) | 1.20 (0.27-5.42) | 2.57 (0.34-19.62) | 1.68 (0.27-10.45) | 1.35 (0.61-2.99) | 1.10 (0.51-2.34) |
| 3<sup>rd</sup> Quartile (0.94, 1.29) | 6.74 (1.70-26.78) | 2.43 (0.58-10.15) | 8.68 (1.40-53.65) | 2.88 (0.49-17.08) | 1.89 (0.87-4.09) | 0.87 (0.37-2.03) |
| 4<sup>th</sup> Quartile (1.30, 1.88) | 10.00 (2.53-39.59) | 4.11 (1.01-16.67) | 13.94 (2.30-84.42) | 5.38 (0.94-30.83) | 2.69 (1.21-5.96) | 1.57 (0.69-3.59) |
| Community population size |  |  |  |  |  |  |
| <10,000 | Reference | Reference | Reference | Reference | Reference | Reference |
| 10,000-499,999 | 1.95 (0.43-8.80) | 1.59 (0.38-6.69) | 2.15 (0.34-13.54) | 1.91 (0.34-10.80) | 2.50 (1.29-4.85) | 2.08 (0.99-4.37) |
| ≥500,000+ | 8.64 (1.99-37.49) | 4.63 (1.05-20.44) | 9.94 (1.67-59.16) | 5.79 (0.98-34.31) | 6.11 (3.00-12.43) | 5.17 (2.06-12.94) |
| **Nursing home factors** |  |  |  |  |  |  |
| Ownership |  |  |  |  |  |  |
| Municipal | Reference | Reference | Reference | Reference | Reference | Reference |
| Private, for-profit | 3.30 (1.72-6.32) | 2.40 (1.14-5.06) | 3.67 (1.68-8.03) | 2.60 (1.04-6.51) | 1.23 (0.71-2.12) | 0.98 (0.54-1.80) |
| Private, non-profit | 2.00 (0.98-4.09) | 2.07 (0.92-4.67) | 2.48 (1.06-5.78) | 2.37 (0.88-6.34) | 1.22 (0.66-2.27) | 1.31 (0.68-2.53) |
| Size of home (beds) |  |  |  |  |  |  |
| <100 | Reference | Reference | Reference | Reference | Reference | Reference |
| ≥100 | 0.83 (0.49-1.40) | 0.90 (0.51-1.59) | 0.70 (0.40-1.22) | 0.74 (0.39-1.39) | 1.77 (1.17-2.69) | 1.48 (0.91-2.41) |
| Crowding Index |  |  |  |  |  |  |
| 1.5 (lowest) | Reference | Reference | Reference | Reference | Reference | Reference |
| 2 | 1.27 (1.14-1.41) | 1.20 (1.04-1.38) | 1.25 (1.11-1.42) | 1.20 (1.01-1.42) | 1.03 (0.91-1.16) | 1.05 (0.90-1.23) |
| 2.5 | 1.61 (1.30-1.99) | 1.43 (1.07-1.92) | 1.57 (1.23-2.01) | 1.44 (1.02-2.02) | 1.06 (0.84-1.36) | 1.11 (0.82-1.51) |
| 3 | 2.05 (1.49-2.82) | 1.72 (1.11-2.65) | 1.97 (1.36-2.84) | 1.72 (1.03-2.86) | 1.10 (0.76-1.58) | 1.17 (0.74-1.86) |
| 3.5 (highest) | 2.60 (1.70-3.98) | 2.06 (1.15-3.67) | 2.47 (1.51-4.02) | 2.06 (1.05-4.07) | 1.13 (0.70-1.84) | 1.24 (0.67-2.29) |

Abbreviations: CI, confidence interval; OR, odds ratio; RR, relative risk

**Figure 3.**
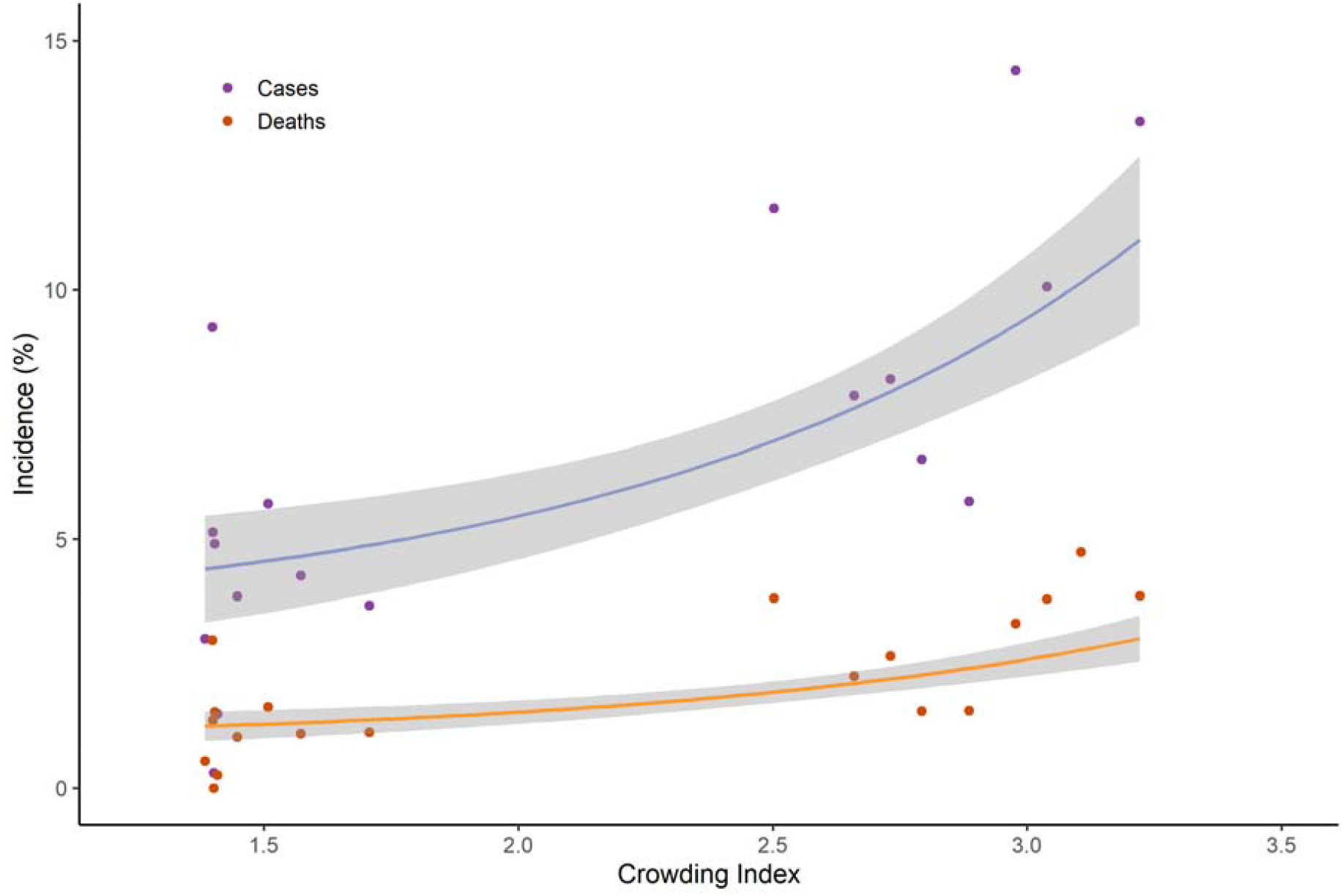
Nursing home crowding index and the incidence of COVID-19 infectionds and deaths, per 100 residents. (N=618 nursing homes). Solid lines represent Poisson regression estimates and shaded regions represent 95% confidence intervals; homes have been aggregated into 20 groups according crowding index, there are 30-31 (5%) of homes in each bin).

In the propensity analysis, a matched home with high crowding index was found for 163 of the low crowding index homes (53%; standardized differences ≤10% for all variables). The resultant effect estimates for incidence (RR=2.06, 95%CI: 1.34-3.17), for mortality (RR=2.09, 95%CI: 1.30-3.38), and for the odds of an outbreak (OR=0.99, 95%CI: 0.58-1.69), were similar to those from regression analyses.

### Preventable Fraction of COVID-19 Incidence and Mortality by Reducing Crowding

Simulation analyses suggested that 988 (18.9%) infections, and 271 (18.7%) deaths in Ontario nursing homes would be prevented if 4-bed rooms had been converted to 2-bed rooms prior to the pandemic. In this scenario, an additional 5,070 new 2-bed rooms would have been needed to maintain capacity across the province, assuming that all current 4-bed rooms were capped at double occupancy. In the simulation where all multiple occupancy rooms were converted to single occupancy, we estimated that 1,624 infections (31.1%) and 450 deaths (31.0%) would have been prevented. In this scenario, an additional 29,871 new single occupancy rooms would have been required, assuming current 4- and 2-bed rooms had been capped at single occupancy.

## Discussion

This study examined whether nursing home crowding was associated with COVID-19 incidence across over 600 nursing homes in Ontario, Canada. The principal findings are that: 1) older nursing homes are more crowded than homes meeting new design standards, and that 25% of residents occupied 4-bed rooms at the onset of the pandemic; 2) COVID-19 infections in nursing homes were extremely concentrated, with almost 90% of infections and deaths occurring in 10% of homes; 3) residents of highly crowded homes were twice as likely to become infected, and die with COVID-19; and 4) simulations suggested that 20% of COVID-19 infections and deaths in Ontario could have been prevented if all 4-bed rooms had been converted to 2-bed rooms prior to the epidemic.

This study provides a comprehensive real-world picture of COVID-19 incidence in 99% of nursing homes in Ontario, Canada. Prior studies examining the impact of nursing home and hospital crowding on respiratory infection spread have largely been limited to controlled before-and-after studies, or smaller cross-sectional studies.^22^ This study leveraged multiple data sources including detailed and standardized nursing home inspections data, provincial data on community COVID-19 infection rates, and census data on community characteristics, in order to provide a population based portrait of crowding and how it impacts COVID-19 incidence, across more than 600 nursing homes.

We found that 26% of Ontario nursing home residents are housed in 4-bed rooms, and that 37% of nursing home residents are housed in 2-bed rooms. This contrasts with preferences among older adults, who prefer single-bed rooms by a margin of 20 to 1, because they confer increased privacy and sense of control.^23,24^ Benefits of multiple occupancy rooms are sometimes touted by nursing home administrators and healthcare professionals, due to lower construction costs, enhanced staffing efficiency, and greater opportunities for socialization.^23,25^ Standards set by many publicly funded healthcare systems only compensate for shared rooms, and as such, private rooms within a nursing home are only available to those able to afford the co-payments; in privately funded healthcare systems, these inequities may be exacerbated.^26^

The crowding index demonstrated that residents of homes with high levels of crowding were twice as likely to develop COVID-19 and die during the first months of the COVID-19 epidemic in Ontario. Our findings align with a prior systematic review that demonstrated that risk of a respiratory infection in occupants of 2-bed rooms was double that of 1-bed room occupants.^22^ Physical barriers such as walls separating bedrooms, strongly predict deposition patterns of viral droplets, much more than absolute distances between beds in a shared room.^27^ Because of this, the crowding index, which is based on persons per bedroom and bathroom, is likely a better predictor than measures based on square footage per occupant.^28^ Risks of COVID-19 due to crowding likely extend beyond just the occupants of crowded rooms, due to the communicable nature of infection and shared spaces in nursing homes.^29,30^ In this case, higher risk among residents of multiple-bed rooms may spillover to residents in single-bed rooms as all residents come into contact with each other either directly, in common areas, or indirectly, via healthcare workers.

Simulation analyses suggested that interventions to de-crowd nursing homes could have prevented approximately 20-30% of COVID-19 infections and deaths. Current guidance from the Centers for Disease Control and Prevention is based on the following pillars: frequent testing of healthcare personnel and residents, facemask use for healthcare personnel and residents, visitor restrictions, supply of hand hygiene and personal protective equipment, and cohorting of known infections and new admissions.^31,32^ But cohorting strategies may be ineffective in crowded homes with many shared rooms. Failure to acknowledge or understand the challenges of cohorting given the overcrowded nature of certain homes, may represent a blind spot of current strategies. This has been specifically noted by nursing home administrators in Ontario.^33^

Insights from this study and the experience of Ontario, can be used to design de-crowding interventions to reduce COVID-19 risk in nursing homes. Such interventions have already been implemented in shelter systems across the United States, wherein shelters have reduced their capacity, and new shelter beds have been added in separate locations.^34^ In Ontario nursing homes, on the basis of early communication of these findings, maximum room occupancy for incoming residents has been capped at 2, as of June 10, 2020.^35^

### Limitations

This study was subject to several limitations. First, the nursing home crowding index and the 1999 design standard were strongly related, and, as such, other design features may have played a role in driving COVID-19 incidence other than crowding.^15^ These could include resident home areas of less than 40 beds (which facilitates cohorting), larger square footage per room occupant, and improved ventilation systems. Second, our examination of crowding was at the nursing home level and we did not know which individual residents occupied single, double, or quadruple occupancy rooms. Third, while we adjusted for aggregate characteristics of nursing home residents, we only had up to date information on nursing home resident characteristics until August 2019, the time of the most recent resident assessment. Fourth, we did not have access to information on nursing home resident race, ethnicity, or socio-economic status.

### Conclusions

Crowding in nursing homes is common and crowded homes are more likely to experience larger and deadlier COVID-19 outbreaks. Interventions to reduce crowding of nursing homes could reduce future COVID-19 incidence and mortality. These interventions could include: 1) capping maximum room occupancy at 2 for incoming residents, to reduce crowding and prevent future outbreaks, 2) creating temporary overflow capacity, to assist with the management of ongoing outbreaks in crowded homes, 3) adapting existing nursing homes (especially converting multiple occupancy rooms into single occupancy) and accelerating the development of nursing homes with exclusively single occupancy rooms, to re-establish nursing home capacity and improve safety for future nursing home residents. Low current occupancy levels due to COVID-19 associated mortality and restricted admissions mean that it may be possible to begin to rapidly and optimally de-crowd entire nursing home systems to prevent further COVID-19 deaths among vulnerable nursing home residents.

## Data Availability

The data from this study is not publically available.

